# Evaluation of Serum Zinc Concentration in Vitiligo, A Case-Control Study

**DOI:** 10.1101/2020.09.16.20195677

**Authors:** Ahmed Abdul-Aziz Ahmed, Hayder Saad Ahmed, Ahmed Talib Abdulqader, Wisam Suhail Najem

## Abstract

**Background:** Vitiligo is an acquired autoimmune pigmentary disorder characterized by ivory white patches. Zinc is proposed to have an immune modulatory, antioxidant and antiapoptotic properties in vitiligo patients. The aim of this study is to evaluate serum zinc in vitiligo patients.

**Patients and methods:** This is case–control study conducted from December 2019 to May 2020. The study included 50 patient with vitiligo and 50 age- and sex-matched controls. A 2 ml venous blood sample was taken from patients and controls by zinc-free syringe and was measured by atomic absorption spectrophotometry.

**Results:** The mean serum zinc level was 73.14±19.32 in patients group and 85.36±14.14 in controls group (p-value=0.005). It also was 77.65±22.07 and 82.84±14.32 in male patients and controls, respectively. In female patients, it was 70.82±17.65and in female controls was 82.88±13.79(p-value=0.02). Regarding the residence, the mean serum zinc level in patients group was 70.13±21.4 in urban and 76.39±17.73 in rural area. In controls group, mean serum zinc was 83.25±14.12 and 89.11±13.77 in urban and rural area, respectively (p-value=0.003). According to the clinical patterns of vitiligo, the mean serum zinc observed was 75.32±19.64 in vitiligo vulgaris, 70.45±21.23 in acrofacial vitiligo and 68.7±17.61 in segmental vitiligo (p-value=0.68).

**Conclusion:** Serum zinc level was significantly low in patients with vitiligo but not associated with the type of vitiligo or family history of autoimmune diseases. It was even lower when vitiligo is associated with nail changes than vitiligo without any nail abnormalities.

## Introduction

Vitiligo is an autoimmune mediated pigmentary skin disorder characterized by well defined ivory white patches that could be localized, acrofacial, segmental or generalized. Melanocytes destruction due to autoimmune, oxidative stress or intrinsic melanocyte defect induced apoptosis represent possible pathogenic mechanisms that ultimately result in total loss of melanin pigment from the skin. The prognosis is variable with unpredictable disease coarse.^1^

The prevalence of vitiligo is variable worldwide. It affects 1% to 2% of population with almost half of patients having a disease onset in childhood. In childhood vitiligo, the females are slightly more affected than males compared to equal gender prevalence among adult patients. ^2^Several autoimmune diseases are associated with vitiligo, particularly thyroid diseases, pernicious anemia, atopic dermatitis, diabetes mellitus and alopecia areata.^3^

Zinc is a trace element that is essential for normal growth and development of different body tissues. It acts as a cofactor for more than 300 metallo-enzymes and 2000 transcription factor required for protein, lipid, nucleic acid synthesis and gene transcription. In addition, it processes an immune modulatory, antiapoptotic and antioxidant effect.^4^ A meta-analysis reported the role of zinc in patients with vitiligo.^5^ The immune modulatory action of zinc, by modulating TNF-a and IL-6, and its antioxidant effect, mediated by superoxide dismutade, protect melanocytes against immune and oxidative stress mediated damage.^6^ In addition, the antiapoptotic effect of zinc mediated by its inhibitory action on caspase-3, caspase-8 and caspase-9 may also play a role in preserving melanocytes.^7^

## Patients and Methods

This is case–control study conducted at Tikrit Teaching Hospital from December 2019 to May 2020. The study included two comparable groups, Patients group involved 50 patient with vitiligo and 50 age- and sex-matched healthy participants as controls group.

Vitiligo was diagnosed clinically by observing macules and patches of depigmented skin which appears as ivory white onWood’s light examination. Only patients and controls who did not use any zinc supplementation in the last month and have no systemic or skin diseases were enrolled in the study.

A 2 ml sample of venous blood was withdrawn from patients and controls by zinc-free syringe and then was heparinized at 40°C. Atomic absorption spectrophotometry was used to measure serum zinc concentration. A 70–120 mg /dl serum zinc was accepted as normal in adults.

The study protocol was approved by theCollege of Medicine, Tikrit University and the local health directorate in Salah Al-Din governorate. In addition, written informed consent provided with complete information about the study protocol was signed by all participants.

Statistical Package for Social Sciences (SPSS) version 22 was used for descriptive statistics calculations. Chi-square, ANOVA and Independent Sample t-test, with a significant P-value of less than 0.05, were used for correlation association assessment.

## Results

The present study included two groups, 50 participants each, as patients group and a comparable controls group. Patients group included 34% (n=17) males and 66% (n=33) females while controls group included 50% (n=25) males and equal number of females (p value=0.4).Urban residence was observed in44% (n=22) and 64% (n=32) compared with rural residence in 56% (n=28) and 36% (n=18) in both patients and controls group, respectively (p value=0.07). The mean age of onset in patients group was 30.86±16.09 and in controls group was 32.22±14.16 (p value=0.6). Table.1

**Table 1:** Demographic data of patients and controls group.

|  | Gender |  |  |  | Residence |  |  |  | Age |
| --- | --- | --- | --- | --- | --- | --- | --- | --- | --- |
|  | Male |  | Female |  | Urban |  | Rural |  |  |
|  | No | % | No | % | No | % | No | % | Mean (SD) |
| Patients group | 17 | 34% | 33 | 66% | 22 | 44% | 28 | 56% | 30.86 (16.09) |
| Control group | 25 | 50% | 25 | 50% | 32 | 64% | 18 | 36% | 32.22 (14.16) |
| P value | 0.4 |  |  |  | 0.07 |  |  |  | 0.6 |

Regarding patients group, the mean age at onset of vitiligo was 22.72±12.29 with the most common observed age at onset was 11-20 years were 34% (n=16) were recorded. There was no significant correlation between mean serum zinc level and age of vitiligo onset (p value=0.07). On the other hand, the mean duration of vitiligo was 7.96±7.30 with most patients 48% (n=24) have disease duration between 1-5 years. The correlation between serum zinc level and disease duration was insignificant (p value=0.1). Table.2

**Table 2:** Age of onset and duration of disease in vitiligo patients group with mean serum zinc level.

|  | Age | No (%) | Serum zinc<br>Mean (SD) | P value |
| --- | --- | --- | --- | --- |
| **Age at onset of vitiligo (years)** | <10 | 10 (20%) | 63.6 (13.87) | 0.07 |
|  | 11–20 | 16 (32%) | 78.25 (17.56) |  |
|  | 21–30 | 10 (20%) | 65.6 (20.27) |  |
|  | 31–40 | 10 (20%) | 74.8 (21.84) |  |
|  | >40 | 4 (8%) | 91.25 (15.37) |  |
| **Duration of vitiligo (years)** | **Duration** | **No (%)** | **Serum zinc**<br>**Mean (SD)** | **P value** |
|  | 1–5 | 24 (48%) | 64.92 (16.93) | 0.1 |
|  | 6–10 | 12 (24%) | 80.42 (20.67) |  |
|  | 11–15 | 10 (20%) | 79 (17.42) |  |
|  | 16–20 | 1 (2%) | - |  |
|  | > 20 | 3 (6%) | 89 (21.16) |  |

Regarding serum zinc level, results showed a mean concentration of73.14±19.32 and 85.36±14.14 in patients and controls group, respectively, (p-value=0.005). Table.3 On the other hand, Table.4 demonstrates serum zinc level in both genders and residence types observed in patients and control group. Mean serum zinc level was 77.65±22.07 and 82.84±14.32in male patients and controls, respectively. In contrast, mean serum zinc level in female patients was70.82±17.65and in female controls was82.88±13.79,(p-value=0.02).Regarding the residence, the mean serum level in patients group was 70.13±21.4 in urban and76.39±17.73 in rural area. In controls group, mean serum zinc was 83.25±14.12 and 89.11±13.77 in urban and rural area, respectively, (p-value=0.003). Table.4

**Table 3:** Mean serum zinc level (mg/ml)in patients and controls group.

|  | No | Mean | SD |
| --- | --- | --- | --- |
| **Patients group** | 50 | 73.14 | 19.32 |
| **Controls group** | 50 | 85.36 | 14.14 |
| **P value** | 0.005 |  |  |

**Table 4:** Mean serum zinc level (mg/ml) in patients and control groups with regard to gender and residence.

|  | Gender |  |  |  | Residence |  |  |  |
| --- | --- | --- | --- | --- | --- | --- | --- | --- |
|  | Male |  | Female |  | Urban |  | Rural |  |
|  | N<br>o | Mean (SD) | N<br>o | Mean (SD) | N<br>o | Mean (SD) | N<br>o | Mean (SD) |
| **Patients group** | 17 | 77.65<br>(22.07) | 33 | 70.82<br>(17.65) | 22 | 70.13 (21.4) | 28 | 76.39<br>(17.73) |
| **Controls group** | 25 | 82.84<br>(14.32) | 25 | 82.88<br>(13.79) | 32 | 83.25<br>(14.12) | 18 | 89.11<br>(13.77) |
| **ANOVA P**<br>**value** | 0.02 |  |  |  | 0.003 |  |  |  |

Different clinical patterns of vitiligo revealed different mean serum zinc level, observed as 75.32±19.64 in vitiligo vulgaris, 70.45±21.23 in acrofacial vitiligo and 68.7±17.61 in segmental vitiligo(p-value=0.68). Only one patient with generalized vitiligo has serum zinc level of 98mg/ml. Table.5

**Table 5:** Mean serum zinc level (mg/ml) in patients with different patterns of vitiligo.

|  | No (%) | Mean | SD |
| --- | --- | --- | --- |
| **Vulgaris** | 28 (56%) | 75.32 | 19.64 |
| **Acrofacial** | 11 (22%) | 70.45 | 21.23 |
| **Generalized** | 1 (2%) | 86 | 0 |
| **Segmental** | 10 (20%) | 68.7 | 17.61 |
| **P value** | 0.68 |  |  |

Family history of associated autoimmune diseases was observed in 70% (n=35) of patients compared with 22% (n=11) in controls group. In decreasing frequency, diabetes mellitus was reported in 38% (n=19) and 6%(n=3), vitiligo in 24% (n=12) and 14% (n=7) and thyroid dysfunction in 8% (n=4) and 2% (n=1) among patients group and controls group, respectively. Differences in serum zinc level between patients and controls group was insignificant (p value>0.05).Table.6

**Table 6:** Family history of autoimmune disorders associated with vitiligo and serum zinc level.

|  | Vitiligo |  | DM |  | Thyroid disease |  |
| --- | --- | --- | --- | --- | --- | --- |
|  | No (%) | Mean (SD) | No (%) | Mean (SD) | No (%) | Mean (SD) |
| **Patients group** | 12 (24%) | 74.58 (19.37) | 19 (38%) | 72.74 (18.38) | 4 (8%) | 60.75 (7.41) |
| **Controls group** | 7 (14%) | 81.14 (12.86) | 3 (6%) | 74 (13.22) | 1 (2%) | 94 (0) |
| **P value** | 0.4 |  | 0.9 |  | — |  |

Nail changes associated with vitiligo were observed in 24% (n=12). Longitudinal leukonychia as observed in 16% (n=8) and leukonychia in 8% (n=4%). Mean serum level was 60.75±12.9and 54.5±11.15 in patients with Longitudinal ridging and leukonychia, respectively, compare to 77.71±18.99in patients with no nail abnormalities (p value= 0.008). No nail changes were observed in controls group. Table.7

**Table 7:** Mean serum zinc level in relation to nail changes in patients with vitiligo.

| Nail findings | Longitudinal ridging |  | Leukonychia |  | Negative |  | P value |
| --- | --- | --- | --- | --- | --- | --- | --- |
|  | No | Mean (SD) | No | Mean (SD) | No | Mean (SD) |  |
| **Patients group** | 8 | 60.75 (12.9) | 4 | 54.5 (11.15) | 38 | 77.71 (18.99) | 0.008 |
| **Controls group** | 0 | 0 | 0 | 0 | 50 | 85.36 (14.14) | - |

## Discussion

Zinc is an essential trace element that may plays a significant role in the pathogenesis of vitiligo. It is a potent free radicals scavenger; thereby reducing oxidative stress mediated destruction of melanocytes. Moreover, the transformation of dopachrome to 5, 6-dihydroxy indole-2 carboxylic acid (DICA) during melanin synthesis is catalyzed by zinc.^6^

In patients group of this study, the serum zinc level was lower than controls, (p-value=0.005). Most studies have demonstrated similar observations. ^8-13^ However, observations made by Doganet al.^14^ 2016, who measuredintraerythrocytes zinc concentration in vitiligo patients, showed no significant differences between patients and controls group. In contrast, Azzam et al.^8^ 2020, who measured serum zinc in 20 patients and equal number of controls, observed higher serum zinc level in vitiligo patients than controls. A systemic review by Huoet al.^15^ 2020 comparing serum zinc level in patients and controls group revealed that higher serum zinc was associated with statistically significant lower risk of vitiligo (OR = 0.47, P < 0.001).

Both males and females in patients group had statistically significant lower mean serum zinc level than controls group, (p-value=0.02). Female patients were affected more than male patients in 66% and 34%, respectively. A study by Sayed et al.^16^ 2015 included 283 patients with vitiligo have also observed a female predominance with193 females (68.2%) and90 males (31.8%), a finding in accordance with our results.

This study also observed that 56% (n=28) of patients were from rural area and 44% (n=22) were from urban area. In addition, mean serum zinc was significantly lower in patients from urban than from rural area, (p-value=0.003). Kuboriet al.^17^ 2006, who reviewed two national surveys enrolling 308 subjects in 2003 and 1017 patients 2005, found that (65%)of subjects were from rural and (44%) were from urban area. Contrary to the present study, a significantly lower mean serum zinc was demonstrated in subjects from rural area compared to urban area. However, non of the subjects involved inKubori et al. review have vitiligo which may explain the contradictory results with our study.

Regarding the vitiligo group, the current study observed no significant correlation between both the age of onset and the duration of disease with mean serum zinc level (p value=0.07 and 0.1, respectively). A case-control study by Basha MA et al.^18^ in 2015 which involved 60 patients and 60 controls observed no significant correlation of serum zinc with age of onset (p value=0.04) or disease duration (p value=0.65). Similar observations were made by Zaki et al.^19^ 2020. In contrast, Mirnezami M et al.^20^ in 2018 found a significantly lower serum zinc level associated with longer vitiligo duration. On the other hand, the present study indicate that the disease duration of 1-5 years was associated with the lower mean serum zinc level, a similar findings was made by Shameer et al.^10^ who observed lower serum zinc level in 2-5 years disease duration.

Vitiligo vulgaris 56% (n=28)is the most common type of vitiligo observed in this study followed by acrofacial 22%(n=11) and segmental vitiligo 20% (n=10), results agree with Handa et al.^21^ 1999 whoalsodemonstratedvitiligo vulgaris as the most common form of vitiligo in69.8% (n=1002), followed by acrofacial vitiligo in14.9% (n=214) and segmental vitiligo in 5% (n=72).Regarding serum zinc level in different pattern of vitiligo, results in this study showed no significant correlation between the clinical type of vitiligo and the measured serum zinc level, (p-value=0.68). These findings were in accordance with Mirza et al.^22^ 2013, who observed no association between serum zinc and different patterns of vitiligo (p value=0.05).

Vitiligo was reported to be associated with a family history of other autoimmune diseases in 70% (n=35) of patients. In decreasing frequency, a family history of diabetes mellitus 38% (n=19), vitiligo 24% (n=12) and thyroid disorders 8% (n=4) were reported in this study; however, with statistically insignificant association with serum zinc level between the two study groups (p value>0.05).A clinico-epidemiological analysis by Khopkar et al.^23^ 2009conducted on 114 patients demonstrated that more than 73% of patients with vitiligo have positive family history of other autoimmune diseases, similar to our observations.

Nail changes were observed in 24% (n=12)of patients group. Longitudinal ridging as the most common finding observed in 16% (n=8) followed by leukonychia in 8% (n=4%). Significantly lower mean serum level was observed in patients with Longitudinal ridging60.75+12.9 and leukonychia 54.5±11.15compared to 77.71±18.99 in patients with no nail abnormalities (p value= 0.008). A case-control study conducted by Topal et al.^24^ 2016 involving 100 patients demonstrated nail changes in 78% of patients. Longitudinal ridging was the most common finding observed in 42% of patients followed by leukonychia, splinter hemorrhage and nail plate thinning. However, no serum zinc level was correlated to nail changes as in the present study.

## Conclusions

Serum zinc level was significantly low in patients with vitiligo but it was not associated with either the clinical type of vitiligo or the family history of autoimmune diseases. Vitiligo associated with nail changes have even lower serum zinc level than vitiligo without nail changes.

## Data Availability

The manuscript was made available on google drive link.

https://docs.google.com/file/d/1V3t798ygFKIePEel6CNSKHv_TzcI0Toe/edit?usp=docslist_api&filetype=msword

## Compliance with Ethical Standards

### Funding

No funding was received from any source.

### Ethical Approval

- All procedures performed were in accordance with the ethical standards of the institutional and/or national research committee and with the 1964 Helsinki declaration and its later amendments or comparable ethical standards.
- This article does not contain any studies with animals performed by any of the authors.

### Informed Consent

Informed consent was obtained from all individual participants included in the study.

